# A one health approach to investigating an outbreak of alimentary tick-borne encephalitis in a non-endemic area in France (Ain, Eastern France): a longitudinal serological study in livestock, detection in ticks, and the first TBE virus isolation and molecular characterization

**DOI:** 10.1101/2021.12.16.21267910

**Authors:** Gaëlle Gonzalez, Laure Bournez, Rayane Amaral Moraes, Marine Dumarest, Clémence Galon, Fabien Vorimore, Maxime Cochin, Antoine Nougairède, Catherine Hennechart-Collette, Sylvie Perelle, Isabelle Leparc-Goffart, Guillaume André Durand, Gilda Grard, Thomas Bénet, Nathalie Danjou, Martine Blanchin, Sandrine A Lacour, Franck Boué, Guillaume Chenut, Catherine Mainguet, Catherine Simon, Laurence Brémont, Stephan Zientara, Sara Moutailler, Sandra Martin-Latil, Nolwenn M Dheilly, Cécile Beck, Sylvie Lecollinet

## Abstract

Tick borne encephalitis virus geographic range and human incidence is increasing throughout Europe, putting a number of non-endemic regions and countries at risk of outbreaks. In spring 2020, there was an outbreak of TBE in Ain, Eastern France, where the virus had never been detected before. All patients but one had consumed traditional unpasteurized raw goat cheese from a local producer. We conducted an investigation in the suspected farm using an integrative One Health approach. Our methodology included (i) the detection of virus in cheese and milk products, (ii) serological testing of all animals in the suspected farm and surrounding farms, (iii) an analysis of the landscape and localisation of wooded area, (iv) the capture of questing ticks and small mammals for virus detection and estimating enzootic hazard, and (v) virus isolation and genome sequencing. This approach allowed us to confirm the alimentary origin of the TBE outbreak and to witness in real time the seroconversion of recently exposed individuals and the excretion of virus in goat milk. In addition, we identified a wooded focus area where and around which there is a risk of TBEV exposure. We provide the first TBEV isolate responsible for as a source of dietary contamination in France, obtained its full-length genome sequence, and found that it does not cluster very closely neither with the isolate circulating in Alsace nor with any other isolate within the European lineage. TBEV is now a notifiable human disease in France, which should facilitate surveillance of TBEV incidence and distribution throughout France.

## Introduction

Tick-borne encephalitis (TBE), caused by tick-borne encephalitis virus (TBEV) is the most important tick-borne zoonotic disease in Europe and Asia from a medical perspective (Süss 2011). Indeed, even though TBEV infection in humans often causes unspecific febrile symptoms and remain un-noticed, it can result in severe neurological diseases including encephalitis, meningitis, and meningoencephalitis, with frequent incomplete recovery and more rarely death (Bogovic, Lotric-Furlan et al. 2010, Kohlmaier, Schweintzger et al. 2021). TBEV is a positive-sense, single-stranded RNA virus of the genus *Flavivirus*, family *Flaviviridae* that circulates preferably among ticks of the genus *Ixodes* and small mammals, but large mammals and migrating birds also contribute to the virus geographic distribution and dispersion (Labuda, Kozuch et al. 1997, Randolph, Miklisová et al. 1999, Waldenström, Lundkvist et al. 2007, Lindquist and Vapalahti 2008, Gunnar, Gunnar et al. 2009, Jaenson, Hjertqvist et al. 2012, Tonteri, Jokelainen et al. 2016, Paulsen, Stuen et al. 2019). TBE results most often from tick bites, however cases can also result from consumption of unpasteurized milk or dairy products from infected cows, goats and ewes (Gresíková, Sekeyová et al. 1975, Holzmann, Aberle et al. 2009, Balogh, Ferenczi et al. 2010, Cisak, Wójcik-Fatla et al. 2010, Caini, Szomor et al. 2012, Hudopisk, Korva et al. 2013, Markovinović, Kosanović Ličina et al. 2016, Brockmann, Oehme et al. 2018, Dorko, Hockicko et al. 2018, Ilic, Barbic et al. 2020, Chitimia-Dobler, Lindau et al. 2021). Indeed, ruminants are also infected when bitten by an infected tick. Even if they remain asymptomatic, virus excretion in milk has been reported for cows, sheep, and goats (Gresíková, Sekeyová et al. 1975, Balogh, Ferenczi et al. 2010, Cisak, Wójcik-Fatla et al. 2010, Caini, Szomor et al. 2012). Pasteurization prevents TBEV transmission through inactivation of virus infectivity (Rónai and Egyed 2020). The increasing geographic range of TBEV, and its patchy distribution around local foci puts an increasing number of countries at risk of sporadic occurrence of TBEV infections that can be both difficult to diagnose and to prevent (Blaskovic 1967, Deviatkin, Kholodilov et al. 2020).

TBEV is endemic throughout Central and Eastern Europe, and has so far been detected in twenty-seven European countries. In the past decades, TBE human incidence has been increasing in several European countries (ECDC 2012, ECDC 2018), with new foci detected in the UK (Holding, Dowall et al. 2020) and Netherlands (Jahfari, de Vries et al. 2017) and TBEV areas expanding in previously unaffected areas (Daniel, Danielová et al. 2004, Lukan, Bullova et al. 2010, Donoso Mantke, Escadafal et al. 2011, Martello, Mannelli et al. 2014, Brabec, Daniel et al. 2017, Rieille, Klaus et al. 2017, Hellenbrand, Kreusch et al. 2019, Alfano, Tagliapietra et al. 2020). These observations associated to evidence of a recent increase of TBEV diversity in Europe suggest that TBEV may be regarded as an emerging disease (Deviatkin, Kholodilov et al. 2020). France is located on the border of the known distribution of TBEV in Europe and very few human cases are usually diagnosed annually, mainly in the Alsace region (about 10-30 cases per year, 0.5/100,000 inhabitants), based on suspect clinical presentation and on the detection of TBEV-specific antibodies (Hansmann, Pierre Gut et al. 2006, Herpe, Schuffenecker et al. 2007, Donoso Mantke, Escadafal et al. 2011, Velay, Solis et al. 2018).

Herein, we report the first outbreak of alimentary TBEV in France and describe the investigation that was conducted within the suspected farm and surrounding area. In April 2020, an outbreak of encephalitis and meningo-encephalitis occurred in the Ain department (Eastern France) where TBEV had never been detected before and a sanitary alert was decreed on May 10. Within a month, epidemiological investigation evidenced that 43 TBE patients with encephalitis, meningo-encephalitis or flu-like symptoms but one had consumed unpasteurized goat cheese or “faisselle” coming from a single local producer (REF TWIN PAPER). The suspected goat flocks were rapidly confined into stall, unpasteurized goat cheese production was stopped, raw cheese produced before the alert were withdrawn from commercial markets, and an investigation was conducted within the farm. We confirmed the alimentary transmission by demonstrating the presence of TBEV in a batch of cheese, and in goat milk. We also monitored the serological status and virus excretion in milk of the animals after control measures had been established, and demonstrated their efficacy, since no TBEV infection have been reported later than early June. We assessed enzootic hazard for animal exposure through tick bite measured as the density of infected ticks in the pasture and in the neighbour woodland. The full-length virus genome sequence was obtained from milk, cheese and tick samples allowing the virus molecular characterization, and an isolate was obtained. Finally, we assessed TBEV seroprevalence in surrounding farms, to evaluate whether TBEV had been silently established in the area or not.

## Material and methods

An overview of the timeline of sampling collections is provided in Figure 1.

**Figure 1.**
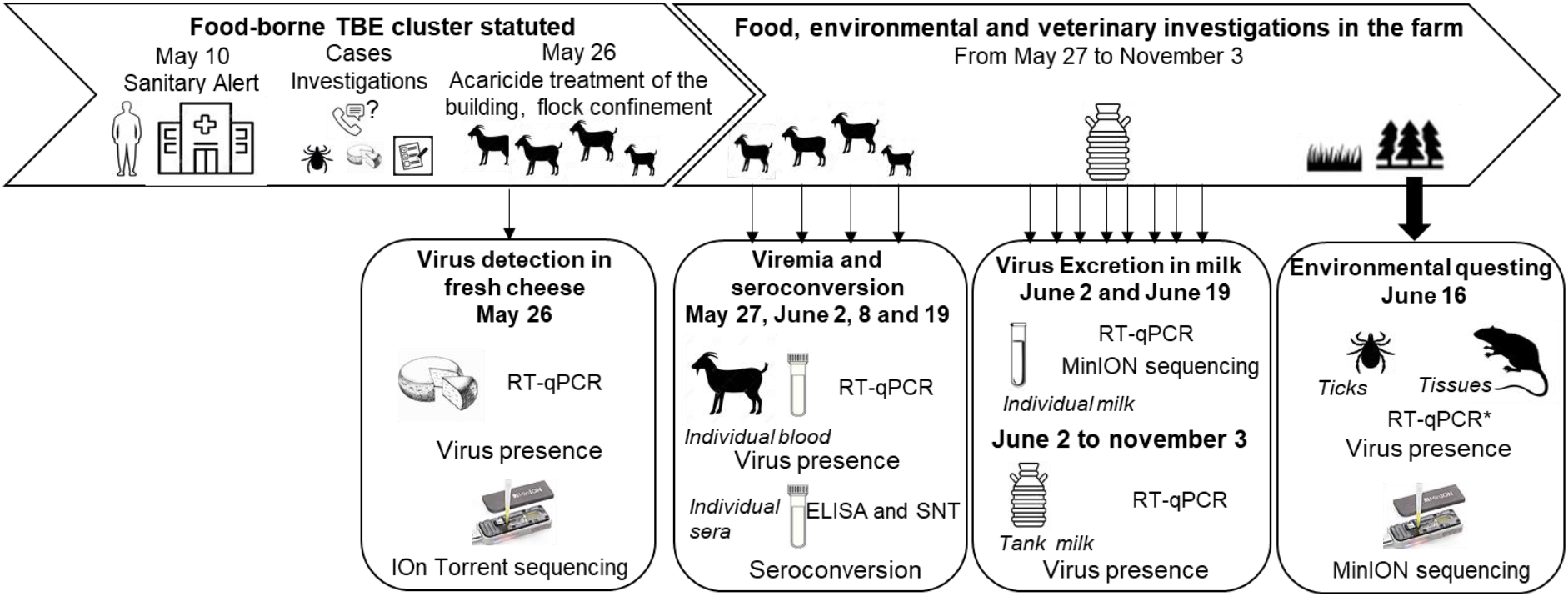
Timeline of the food, environmental and veterinary investigations and sampling scheme, April-November 2020

### Cheese sampling

Cases investigations performed by the French public health Agency pointed out that one case still had cheese from the suspected farm at home at the time of the investigation. This fresh cheese was produced on April 28. The Ain local authorities recalled cheese produced at the suspected farm. Five samples from the batch produced on April 28 and 74 from other batches were sent to the National Reference Centre (NRC) for Arboviruses for screening of TBE genome. Confirmation was conducted at the Food-safety national reference laboratory, ANSES, Maisons-Alfort.

### Serum, blood, milk and tick sampling

#### From animals at the suspected farm

The farm suspected to be the source of the infection included 56 dairy goats, three dairy cows and four suckling cows grazing on the same pasture. When the epidemiological investigation led to determine that this farm was strongly suspected to be the source of the human contamination, all goats and dairy cows were confined into the stall and cows were treated against ticks on May 26, 2020. Acaricide treatment of the flock buildings was also carried out. Serum and blood samples were collected as follows (Figure 1). On May 27, serum samples were collected from 30 goats. On June 2, 8 and 19, serum and blood samples were collected from all 56 goats of the farms. Serum samples were collected on the three dairy cows on June 2 and on the four suckling cows between July and August 2020. In addition, tank milk samples from goats and dairy cows were collected every week from June 2 to November 3. Individual milk samples were collected from the 55 lactating goats on June 8 and 19. In March 2021, a follow-up serological study was carried out on the entire goat herd among which 35 individuals were identical to 2020. Milk and sera were stored at 4°C and blood samples were stored at -20°C until use.

#### From animals in surrounding farms

In order to characterize the geographic distribution of the TBEV infections among the surrounding farms, one hundred and forty-two animals from five farms – two goat farms, one dairy cow farm and two suckling cow farms – from which the animals have grazed in pastures located less than 2 km away from the infected farm’s pasture were investigated for the presence of TBEV antibodies. Serum samples of all animals available at the time of sampling were collected between June and August 2020.

#### Collect of questing ticks

Questing ticks were counted and collected from 10 a.m. to 6 p.m on June 16, 2020 by dragging a 1-m^2^ white blanket. The pasture was divided into 5 geographic areas in which a total of 29 sub-areas was defined according to the habitat types: wooded areas (3 sub-areas), wooded edge of the meadow (meadow-forest or meadow-wood ecotones, 5 sub-areas), hedgerows along the edge of the pasture or within the pasture (7 sub-areas), meadow between 3 and 10 meters away from the forest edge (4 sub-areas) and from the hedgerows (4 sub-areas), meadow more than 10 meter from the edge (6 sub-areas). The adjacent forest was divided into 3 sub-areas. The sampling scheme differed according to the sub-areas, with more effort made in wooded areas in order to collect more ticks for TBEV analysis. A total of twenty-eight 10-meters-long transects were carried-out in either forest (18 transects) and wood (10 transects) sub-areas. In addition, the edge of the meadow and hedgerows was continuously checked for tick presence and individuals were collected every 20-meters-long transects. Within the meadow, five 20-meters-long transects were realized. Because the dragging method is more appropriate for estimating the density of questing nymphs than those of larvae and adults, only the density of questing nymph per sub-area was estimated by evaluating the number of ticks collected per 100m^2^ and its mean per habitat type was calculated.

We calculated that, for an estimated virus prevalence in nymphs of 0.2%, it would be necessary to collect and test for TBEV presence in a minimum of 1000 nymphs in order to have a 95% chance to detect the virus (Cannon 2001). Given that previous estimates of TBEV prevalence in the French endemic region (Alsace) ranged from 0.03 to 0.3% in nymphs, and from 0.6 to 0.8% in adults (Perez-Eid, Hannoun et al. 1992, Bestehorn, Weigold et al. 2018, Bournez, Umhang et al. 2020), we conducted an additional collect of ticks in the forest to reach that critical minimum number. Back in the laboratory, ticks were identified to the genus level based on their morphology (Pérez Eid 2007). All questing nymphs and adults were washed in 70% ethanol, rinsed twice in distilled water, dried, and stored at ™80°C until used for TBEV detection.

#### Small mammal trapping and sampling of feeding ticks

For three consecutive nights from June 15 to 18, 2020, two hundred small mammal live-traps (14 × 14 Ugglan special no. 3, Grahnab, Sweden) were set up every 10 meters along the pasture and within the wooded area of the pasture. Traps were baited with carrots and sunflower seeds. Caught animals were euthanized and identified at the genus level. Spleen and brain of small mammals were collected and put in nitrogen container until storage at -80°C in the laboratory.

All efforts were made to minimize animal suffering. The species captured during this study are not protected in France nor included in the International Union for Conservation Nature Red List of threatened species and, therefore, no special authorization was needed for trapping, according to French law. Animal trapping took place with permission from the landowners.

Ticks feeding on trapped animals were removed and identified to the genus level based on their morphology (Pérez Eid 2007). Potential undetected ticks were collected by removing the skin of the animals and storing them in individual bags for 48 hours to let the ticks detach.

### Laboratory analyses

#### Serological tests

Antibodies against flaviviruses were detected using a multi-species commercial competitive enzyme-linked immunosorbent assay (cELISA; ID Screen^®^ West Nile Competition, ID Vet, Montpellier, France) based on purified whole WNV antigen for the detection of antibodies directed against the envelope E protein common to all flaviviruses. Although this ELISA test is designed for WNV detection, it can be used for the detection of TBEV antibodies because of the high cross-reactivity among flaviviruses (Rushton, Lecollinet et al. 2013, Beck, Desprès et al. 2015, Beck, Fritz et al. 2016, Bournez, Umhang et al. 2019, Reusken, Boonstra et al. 2019). The test was performed according to the manufacturer’s instructions. The interpretation was modified for sera close to the doubtful and negative thresholds, with extension of the doubtful interval, to ascertain the detection of sera with low-TBEV antibody levels by ELISA and then increase sensitivity. The sample was considered positive if < 40%; doubtful between 40% and 72% and negative if > 72%.

In-house IgM-capture enzyme immunoassay (MAC-ELISA) with whole inactivated TBEV was performed as previously described (Peyrefitte, Pastorino et al. 2005) on all 30 goat sera sampled on May 27, 2020. MAC-ELISA was performed using rabbit anti-goat IgM antibodies (Bethyl Laboratories, Inc, USA).

Samples with positive and doubtful results in ELISA were then tested for the presence of specific neutralizing antibodies against TBEV by micro virus neutralization tests (MNTs, strain Hypr, Genbank ID U39292.1) as described in Beck et al. (2015) (Beck, Desprès et al. 2015). A serum sample was considered positive for TBEV if cells were protected at least at the serum dilution 1:20.

#### RNA extraction and real-time TBEV qRT-PCR

##### qRT-PCR tests on fresh cheese

Cheese suspected to be infected with TBEV were stored at -80°C until analysis. The National Reference Centre for Arboviruses carried out TBEV genome screening following the protocol bellow (REF TWIN paper). Dissolution of nearly 5g of fresh or drier cheese was performed in PBS. Homogenization steps were achieved to complete the process. RNA extraction was carried out using the QIAmp Viral RNA Mini Kit (Qiagen, Paris, France) according to manufacturer instructions. Five microliters of the eluted RNA were used to execute a quantitative RT-PCR with primers and probe targeting TBEV genome (sequences available on request) using the SuperScript™ III Platinum™ One-Step qRT-PCR amplification kit (Thermofischer, Paris, France) on LightCycler® 2.0 Instrument (Roche Life Science, Germany).

Confirmation of TBEV-infected cheese was conducted at the Food-safety national reference laboratory, ANSES, Maisons-Alfort. The method used to recover TBEV from fresh cheese was based on the use of proteinase K as previously described in Martin-Latil et al. (2017) (Hennechart-Collette, Martin-Latil et al. 2017). After enzymatic digestion, total nucleic acid extraction was carried out using the NucliSENS® easyMAG™ platform, according to the manufacturer’s instructions (bioMérieux). Primers and probe used to detect TBEV by RT-qPCR have been already described in the literature by Gondard et al (2018).

#### qRT-PCR tests on goats, cows and small mammal samples

Goat and dairy cows tank milks, goat individual milk samples, goat or cows EDTA blood, and the spleen and brain of small mammals suspected to be infected with TBEV were stored at 4°C (milk), -20°C (EDTA blood) or -80°C (organs) until analysis. RNA extraction was performed using the MagVet™ Universal Isolation kit (Lifetechnologies, France) on the King Fisher automat (ThermoFisher Scientific, France). Five microliters of each RNA extracts were subjected to qRT-PCR with primers and probe targeting the NS5 gene described in (Gondard, Michelet et al. 2018) using the AgPath-ID™One-Step RT-PCR Reagents kit (Lifetechnologies) and cycling conditions as followed: reverse transcription for 10 min, 45°C; denaturation of cDNA 10 min at 95°C and 45 cycles of 15 sec at 95°C and 1 min at 60°C. The detection limit of the qRT-PCR, that is the lowest number of copy genome detected for a known dilution in 95% of cases, performed on goat milk samples was estimated at 1.10e4 TCID50/ml corresponding to a Ct value of 34.

#### qRT-PCR tests on questing ticks and descriptive analysis of TBEV infection in ticks

Questing ticks were analysed to detect TBEV RNA. Adult ticks were analysed individually and nymphal ticks were analysed in pools of 30 ticks maximum per sub-area. RNA were extracted using the Nucleospin RNA II extract kit (Macherey-Nagel, Düren, Germany) as described in (Bournez et al., 2020) and were screened for TBEV by q RT-PCR targeting the NS5 as described in (Gondard, Michelet et al. 2018). Because TBEV infection prevalence in ticks is lower than 1% in endemic regions of France, prevalence in nymphs was expressed as the minimum infection rate per 100 tested (MIR), based on the assumption that a single tick was positive within a positive pool. We calculated the overall MIR of TBEV in nymphs and TBEV prevalence infection in adults and their 95% CIs were calculated following a binomial distribution. We estimated the density of infected nymphs per habitat by multiplying the % MIR by the mean density of questing nymphs per habitat.

#### Virus isolation from contaminated milk and infected ticks

Contaminated milk (500 µL) or tick grindings (100 µL/homogenates) positive for TBEV were diluted in serum-free DMEM culture medium and inoculated in a T25 flask seeded with Vero NK cells (ATCC: CCL81™) 24 h earlier, and washed with DMEM before inoculation. Following an incubation time of 1h30 at 37°C with 5% CO2, cells were washed twice with phosphate buffered saline (PBS), and complete medium (DMEM+ 1% penicillin-streptomycin+ 1% sodium pyruvate + 5% fetal calf serum) was added. The cells were observed every day from day 3 to day 7 post-infection (pi). As soon as cytopathic effects (CPE) were detected, the supernatant was collected, clarified and stored at −80 °C. RNA extraction was performed using the MagVet™ Universal Isolation kit (Lifetechnologies, France) on the King Fisher automat (ThermoFisher Scientific, Paris, France). RNA extracts were subjected to TBEV RT-qPCR as described above to confirm TBEV detection.

#### Full-length TBEV genome assembly performed from contaminated milk and infected ticks

Full-length virus genome sequencing was conducted on cDNA obtained from contaminated milk and infected ticks. Multiplex primers were designed using Primal scheme (http://primal.zibraproject.org) with default parameters. A multifasta file with reference genomes NC_001672.1, KF151173.1, MG589938.1, KC835596.1, MK801808.1, MK801803.1, MG210948.1, KX966399.1 and KP716974.1 was used to generate primers. Their sequences are listed in Table 1 (Supplementary data) (scheme.primer.tsv). A multiplex PCR method for targeted enrichment was adapted for library preparation and MinION sequencing of TBEV genome as previously described for Zika (WHO Control Reference 11474/16) and Chikungunya viruses (PEI N11602) in (Quick, Grubaugh et al. 2017).

**Table 1.** Summary of the test results carried out on serum and milk samples collected from the goats of the suspected farm. Serum samples were initially tested by a competitive ELISA, then positive and doubtful ELISA samples were tested by specific-TBEV virus micro-neutralization tests (MNT). A serum sample was considered TBEV-seropositive when MNT was positive. *Two goats were MNT negative at one of these two dates but considered TBEV positive because clearly positive by MNT on 2020/06/02 (see supplementary data). Moreover, both of them excreted TBEV in the milk.

| Date | Serum<br>tested by serological tests |  |  | Individual milk<br>tested by qRT-PCR |  |  | Tank milk<br>tested by RT-PCR |  |
| --- | --- | --- | --- | --- | --- | --- | --- | --- |
|  | No. | No. TBEV<br>positive | %<br>TBEV-positive | No | No.<br>TBEV<br>positive | %<br>TBEV-positive | yes/no | Ct value |
| 27/05 | 30 | 6 | 20 | 0 |  |  | no |  |
| 02/06 | 56 | 11 | 20 | 0 |  |  | yes | 35 |
| 09/06 | 56 | 13* | 23 | 55 | 3 | 5.5 | yes | neg |
| 23/06 | 56 | 13* | 23 | 55 | 0 | 0 | yes | neg |
| 07/07 | 0 |  |  | 0 |  |  | yes | neg |
| 15/07 | 0 |  |  | 0 |  |  | yes | neg |
| 28/07 | 0 |  |  | 0 |  |  | yes | neg |
| 04/08 | 0 |  |  | 0 |  |  | yes | neg |
| 11/08-03/11 | 0 |  |  | 0 |  |  | yes | neg |
| 2021 | 53 | 8 | 15 | 0 |  |  | no |  |

Sample barcoding was performed with 14 barcodes (NB01 to 14) with the native barcoding genomic DNA protocol using the ligation sequencing kit SQK-LSK109 (Oxford Nanopore technologies, Oxford, UK) and the Native barcoding expansion kits EXP-NBD104 and EXP-NBD114 (Oxford Nanopore technologies, Oxford, UK). Two barcodes were assigned per sample, depending on the pool of primers used (either pool 1 or 2) (Supplementary data Table 2). A Flongle flow cell FLO-FLG001 was used for sequencing (Oxford Nanopore technologies, Oxford, UK).

FAST5 reads were base-called offline using guppy_basecaller v5.0.7. from Oxford Nanopore (https://nanoporetech.com). Subsequent FASTQ files were demultiplexed using guppy_barcoder using the --trim-barcodes flag. 25 nucleotides were trimmed at both 5’ and 3’ ends in resulting FASTQ files to remove primers and with a maximum length of 700 bp using NanoFilt v2.6. The sequences obtained with the two barcodes corresponding to a unique sample were concatenated into one FASTQ file. Each file were mapped to the reference NC_001672.1 using minimap2 v2.20 (Li 2018). Consensus FASTA files were generated after 4 error correction steps including three polishing steps of racon v1.4.20 (https://github.com/isovic/racon) and one polishing step with medaka v1.4.3 (https://github.com/nanoporetech/medaka). Trimmed reads were mapped back to the assembly using minimap2 and coverage statistics were calculated on the sorted BAM file using SAMtools v1.12 (Li and Durbin 2009). Multiple alignments of the nucleotide sequences were performed using the ClustalW algorithm to extract a high quality full-length genome sequence.

#### Full-length TBEV genome assembly performed from contaminated cheese

Full-length virus genome sequencing was conducted on cDNA obtained from contaminated cheese using the ProtoScript® II Reverse Transcriptase kit (New England Biolabs) with 50µM random hexamers (Invitrogen) (7:1 ratio) for 5 minutes at 65°C (Bournez, Umhang et al. 2020). The full-length genome was amplified (multiplex PCR) with Q5® High-Fidelity 2X Master Mix kit (New England Biolabs) and a set of primers designed using the ‘Primal Scheme’ program (http://primal.zibraproject.org/) (Quick, Grubaugh et al. 2017). The list of primers used for sequencing is presented in supplemental table 3. Sequencing analysis was performed using the S5 Ion torrent technology v5.12 (ThermoFisher Scientific) as previously described (Driouich, Moureau et al. 2019). The consensus sequence was obtained after a trimming step of reads (reads with quality score < 0.99, and length < 100bp were removed and the thirty first and thirty last nucleotides were removed from the reads). Mapping of reads to a reference (determined following blast of De Novo contigs) was realized using CLC genomics workbench software v.20 (Qiagen). A *de novo* contig was also generated to ensure that the consensus sequence was not impaired by the reference sequence. For genomic regions for which no sequence was obtained using this approach, cDNA was also generated and amplified using the SuperScript™ IV One-Step RT-PCR System kit following the supplier’s recommendations (Invitrogen). PCR mixes (final volume 50µl) contained 3µl of cDNA, 2.5µl of each primer (final concentration of 0.5µM), and 25µl of 2X Platinum™ SuperFi™ RT-PCR Master Mix. Amplification was performed with the following conditions: 50s at 50°C, 2min at 98°C, then 40 cycles of amplification (10s at 98°C, 10s at 66°C, 30s at 72°C) and a final extension of 5min at 72°C. PCR products were verified by gel electrophoresis and pooled. Sequencing analysis was performed as described above. The consensus sequence was obtained using the reads from both runs as described above.

#### Phylogenetic analyses

The novel reference genome sequence was aligned against the known TBEV viral diversity. Nucleotide and protein percentage of identity were calculated with Clustal Omega (Madeira, Park et al. 2019). Phylogenetic analysis were conducted on the full-length open reading frame. Phylogenetic tree were then inferred using the maximum likelihood method implemented in PhyML (version 3.0) (Lefort, Longueville et al. 2017) using the best-fit model and best of NNI and Subtree Pruning and Regrafting (SPR) branch swapping. Support for nodes on the trees were assessed using an approximate likelihood ratio test (aLRT) with the Shimodaira-Hasegawa-like procedure. Trees generated using the Neighbor– Joining and Maximum Likelihood methods implemented in MEGAX gave identical results.

## RESULTS

### TBEV detection in fresh cheese produced in the suspected farm

On June 2, the NRC Arbovirus detected one cheese TBEV positive over seventeen (REF TWIN paper). This cheese produced on April 28 was collected on May 26 in a patient home during the investigation time. The local authorities suspected a food-born source of contamination as patients developed meningo-encephalitis and all consumed goat cheese produced in the same farm. The presence of TBEV genome was confirmed by the ANSES food-safety laboratory (Maisons-Alfort). The food-borne origin of TBEV contamination was therefore confirmed. On July 10, seventy-four cheese given by the producer were analysed and virus genome was detected in six of them.

### TBEV infection and exposure in suspected farm livestock

IgG and IgM antibodies against TBEV were detected using ELISA tests (Igs competition and IgM capture), and MNT. On May 27, six goats over thirty were TBEV-seropositive as assessed with competition ELISA and MNT. One of them had anti-TBEV IgM antibodies. The following weeks, after their placement in a stable to prevent further tick bites, all 56 goats were tested for a complete assessment of TBEV exposure rate. Eleven goats were TBEV-seropositive on June 2 (20%) (with one seroconversion in the 30 goats collected the 27th of May) and 13 (23%) on June 9 (with two more seroconversions) (Figure 2). Goat seroconversions were confirmed by MNT (Figure 2, Table 1 and supplementary data). Overall, ELISA and MNT results agreed, with the exception of a single individual that appeared positive based on ELISA competition but did not develop neutralizing antibodies (Figure 2). The seroconversion of three goats between May 27 and June 9 indicates that infection occurred very recently. Yet, none of the animals had high viremia given that all qRT-PCRs conducted on blood samples were negative (collected on June 2, 9 and 19). ELISA competition conducted in 2021, about one year after the initial infection, showed a decrease in IgG levels for most seropositive individuals (10/13) but the level of neutralizing antibodies remained high. Two individuals that were TBEV-seropositive in 2020 turned negative based on competition ELISA in 2021, and one of those had no remaining neutralizing antibodies according to MNT (Figure 2).

**Figure 2:**
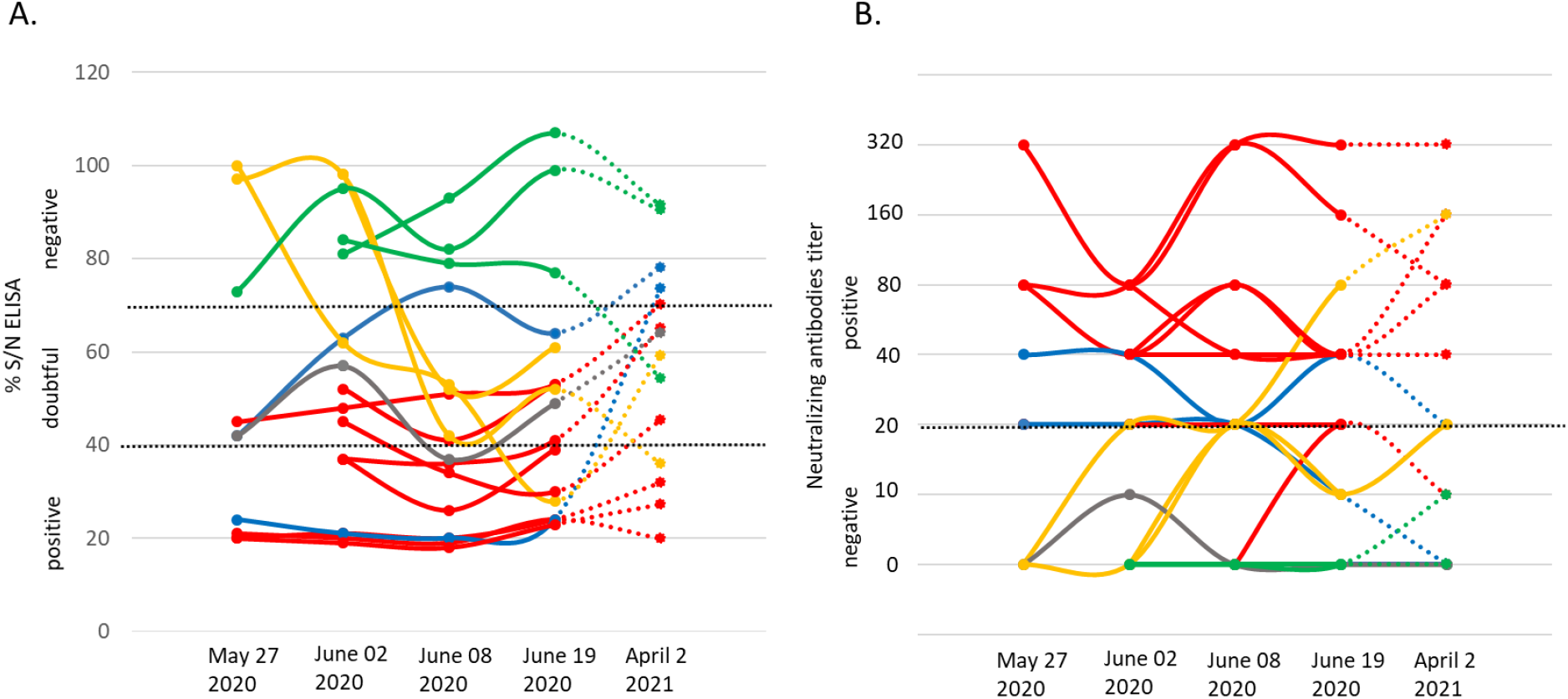
IgG levels (A) and micro-neutralization antibody levels (B) in goats on four dates in 2020 (May 27, June 02, June 08 and June 19) and on April 2, 2021. The threshold levels applied are indicated as dashed lines. We provide examples of seronegative individuals (green), and show results for the individual that returned seropositive via ELISA competition, but returned negative neutralizing antibody titre (grey), the three individuals that seroconverted after May 27 (yellow), the eight seropositive individuals (red) and the two individuals that returned negative IgG competition results in 2021 (blue), one of which maintains neutralizing antibodies.

Within the same farm, one of the three dairy cows and two on the four suckling cows, which had grazed in the same or in the adjacent pasture as the goats, were TBEV seropositive. No virus genome was detected in the blood of these animals.

### TBEV detection in goat milk at the suspected farm

The presence of TBEV genome was detected in the tank of goat milk collected on June 2, but it was absent in the tank of cow milk. The virus was not detected in either tanks thereafter (between June 8 and November 3). The presence of TBEV was tested in the milk of individual goats collected on June 8 and June 19. TBEV genome was detected in the milk of three goats on June 8 (23% of the thirteen seropositive animals) indicating that they continued excreting viruses 14 days after their confinement; two individuals were TBEV-seropositive on May 27 and one seroconverted between June 2 and 8. On June 19, all individual milk samples returned negative results.

### Seroprevalence survey in surrounding farms

Within a couple of kilometres away from the suspected farm, five farms, hosting cows and goats were identified and sera from all individuals were collected and processed as above. Few seropositive animals were found in three of the five farms, with one to five seropositive animals and a TBEV seroprevalence ranging from 5.5% to 25% (Figure 2, Table 2). Animals from the suspected farm, and from farms #1 and #3 within which seropositive animals were found, have all grazed in meadows located in close proximity to the same forest (Figure YY). In farm #4, a single 10 year-old cow was TBEV-seropositive and it was not possible to trace back where the cow had grazed in the past.

**Table 2.** Number of questing ticks collected and mean density per habitat.

| Habitat | No. sub-areas | Sampled surface (m<sup>2</sup>) | No. nymphs | No. female | No. male | Density of nymphs (/100 m<sup>2</sup>) |  |
| --- | --- | --- | --- | --- | --- | --- | --- |
|  |  |  |  |  |  | mean | Range |
| Forest | 3 | 540 | 382 | 10 | 20 | 71.5 | 64.2-81.3 |
| *additional collection* | / | / | 285 | 7 | 5 | / | / |
| Pasture |  |  |  |  |  |  |  |
| - wooded area | 3 | 300 | 105 | 6 | 3 | 31.7 | 20.0-50.0 |
| - wooded edge | 5 | 400 | 94 | 2 | 2 | 24.0 | 5-56.7 |
| - hedgerows | 7 | 870 | 4 | 0 | 0 | 0.3 | 0-1.1 |
| - meadow between 3 to 10m from the wooded edge | 4 | 530 | 35 | 2 | 1 | 4.5 | 0-8.8 |
| - meadow between 3 to 10m from the hedgerow | 4 | 400 | 2 | 0 | 0 | 0.3 | 0-1.1 |
| - meadow >10m from the edge | 6 | 600 | 0 | 0 | 0 | 0 | 0 |

### TBEV presence in ticks and rodents within and around the pasture

The goats have grazed in only one pasture adjacent to the dairy cow farm. Half of the goat pasture was a wooded area, which was in continuity of a large mixed deciduous and coniferous forest dominated by beech trees.

#### Detection of TBEV in ticks

A total of 120 larvae, 907 nymphs, 27 females and 31 males of questing *Ixodes* spp. and 1 female of *Dermacentor reticulatus* were collected within the pasture and in the adjacent forest. Within the pasture, ticks were only found in the wooded area, along the wooded edge and hedgerow and in the meadow situated less than 10 m away from the forest edge. The density of questing *Ixodes* spp. nymphs was the highest in the forest with a mean of 71.5 nymphs/100 m^2^, then in the wooded areas and in the wooded edge of the pasture with a mean of 24-32 nymphs/100 m^2^ and finally in the meadow with a mean of 4.5 nymphs/100 m^2^ (Table 2).

Two pool of nymphs -from the forest and from the meadow located between the wooded area of the pasture and the forest - and one male from the forest were subsequently tested positive for TBEV RNA. Overall, the MIR was 0.22% (IC_95%_ : 0.03 – 0.80%) in nymphs and the infection rate was 1.8% (IC_95%_ : 0.1 – 9,9%) in adults. The estimated density of infected nymphs was the highest in the forest with 0.15 infected nymphs / 100 m^2^, then in the wooded area and in the wooded edge of the pasture with 0.05- 0.06 infected nymphs / 100 m^2^.

#### Absence of TBEV in captured small mammals

In addition, small mammals were caught around the pasture to assess TBEV presence. In total, ten small mammals were captured: nine shrews (*Sorex* spp.) and one bank vole (*Myodes glareolus*). Only the bank vole was captured at the edge of the forest. Larvae of *Ixodes ricinus* were observed respectively on the bank vole (five larvae) and on two shrews (two larvae). All animal organs (spleen and brain) were negative to TBEV as assessed by qRT-PCR.

### Phylogenetic analysis

Three RNA samples extracted from two TBEV-positive pools of nymphs and one TBEV-positive male tick and RNA samples obtained from three individual infected-goat milks were subjected to sequencing. Our amplicon-based sequencing protocol allowed us to successfully sequence TBEV genome from RNA extracted from milk and ticks (Figure 4). The full-length genome sequence obtained from milk and ticks was similar to the full-length genome obtained from the contaminated cheese collected during the investigation led by local authority agency. Sequences obtained from the milk sample and the virus isolate obtained from ticks were 100% identical except for single nucleotide insertions and deletions that are most likely due to sequencing errors.

**Figure 3.**
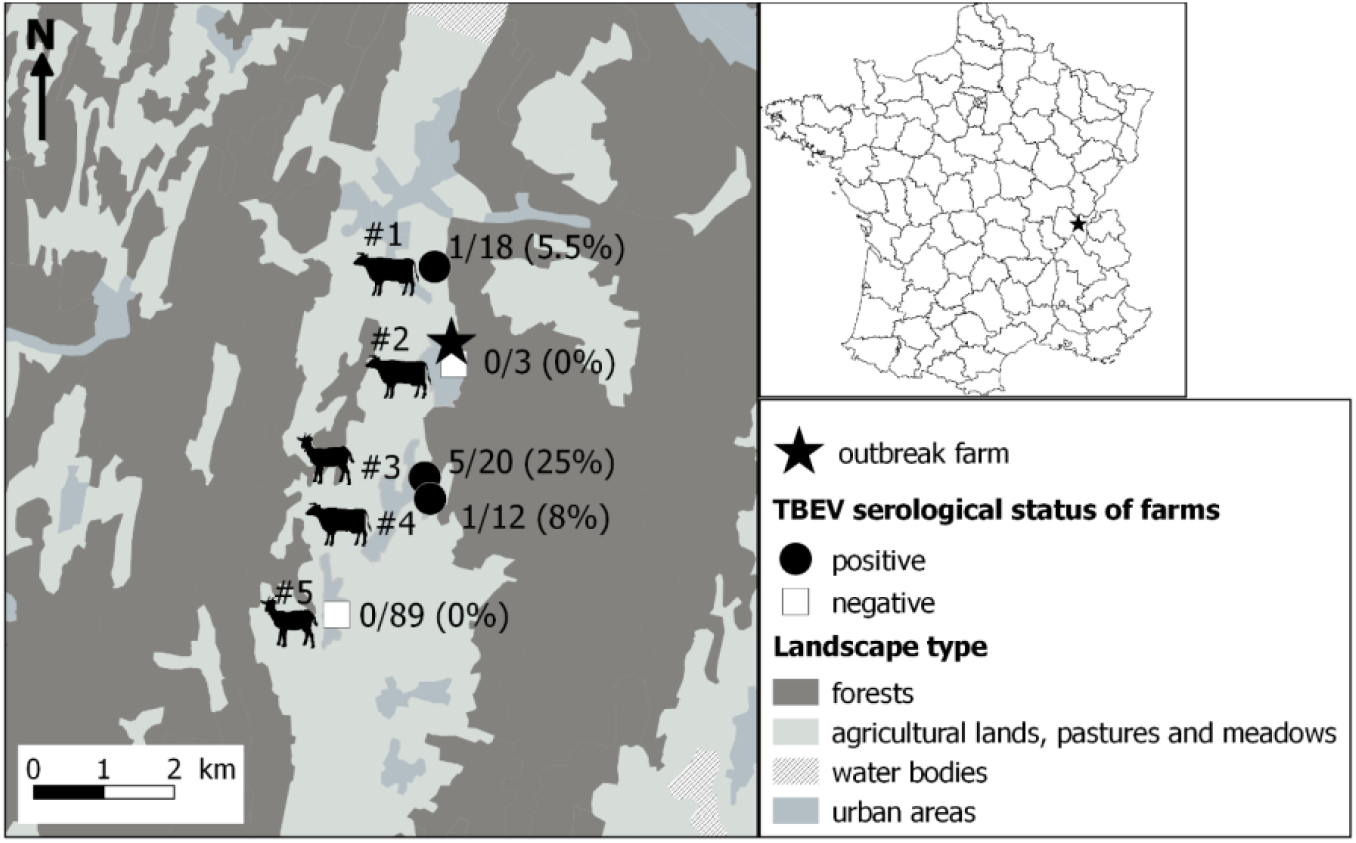
Localisation of the farm responsible for the tick-borne encephalitis cluster through alimentary route (star) and results of the serological survey conducted in surrounding farms. Serum sample were first tested by a competitive ELISA, then positive and doubtful ELISA samples were tested by specific-TBEV virus micro-neutralization tests (MNT). A serum sample was considered TBEV-seropositive when MNT was positive. Dairy cows, suckling cows, goats, suckling cows and goats were found in farms #1 to #5, respectively.

**Figure 4:**
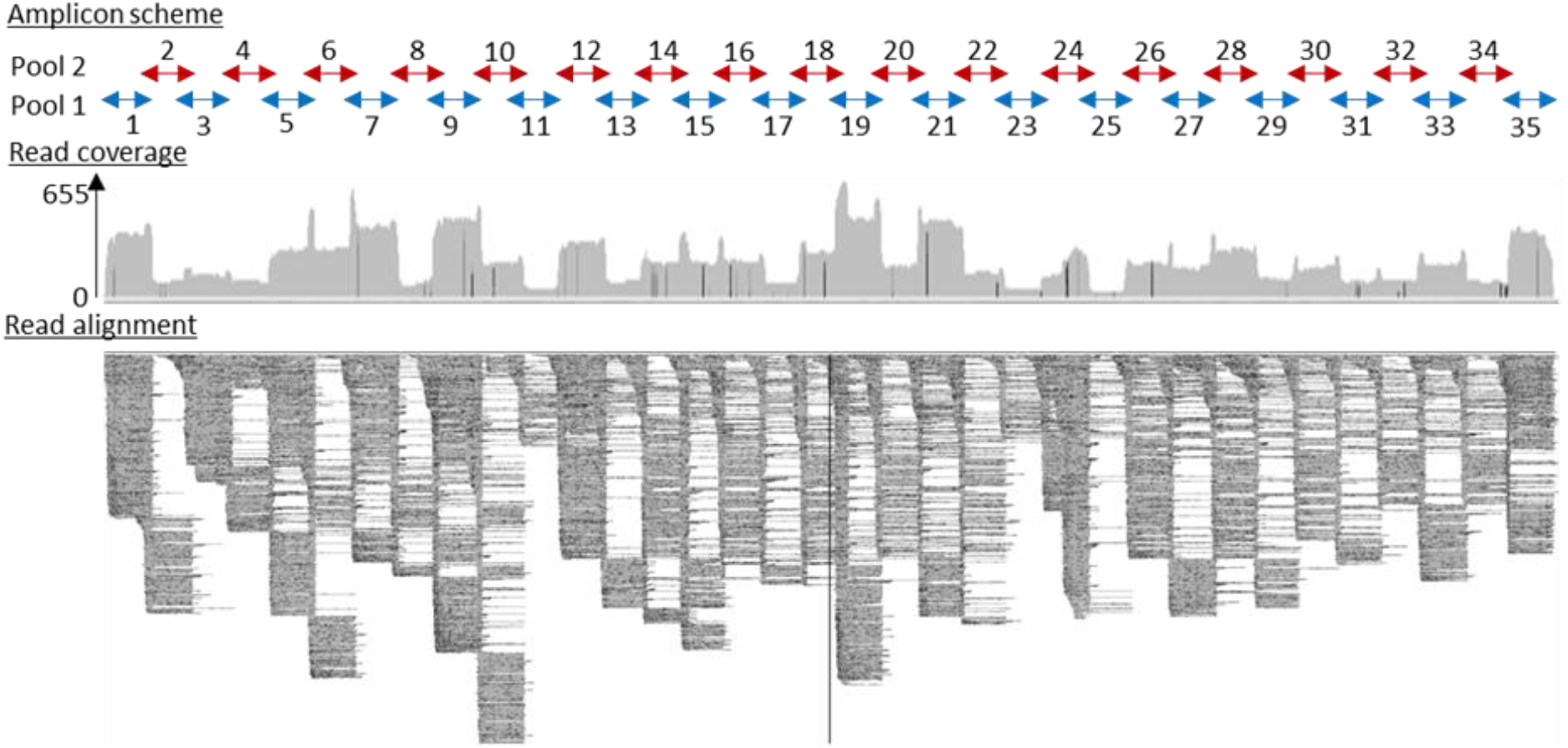
Full-length TBEV genome sequence (GenBank accession number: OL441148) was obtained from sequencing 35 overlapping amplicons in two multiplex reactions. Read coverage varied along the sequence length, but a full-length genome sequence was successfully recovered.

The full-length genome of the new viral strain was sequenced and compared to previously characterized TBEV genome sequences. The TBEV_Ain_France_2020 had limited homology (98.4% identity) to the partial NS5 of the TBEV from the Alsace region (AF0910) (Supplementary data). The nucleotide sequence presents 98.8% identity to the same partial NS5 and between 98 and 98.07% identity to the full-length genomes of TBEV isolates detected in Russia, Germany, Austria, Czech Republic and the Netherlands with the most ancient isolate dating from 1951 in Russia. The polyprotein sequence presented up to 99.30 % and 99.24% identity to MK922616 and MK922617 isolated from ticks collected in the German federal state of Lower Saxony in 2018. Our phylogenetic analyses confirmed that TBEV_Ain_2020 (OL441148) belongs to the European subtype (TBEV-Eu3) and is most closely related to TBEV strains recently isolated in bordering countries and Eastern Europe (Figure 5).

**Figure 5:**
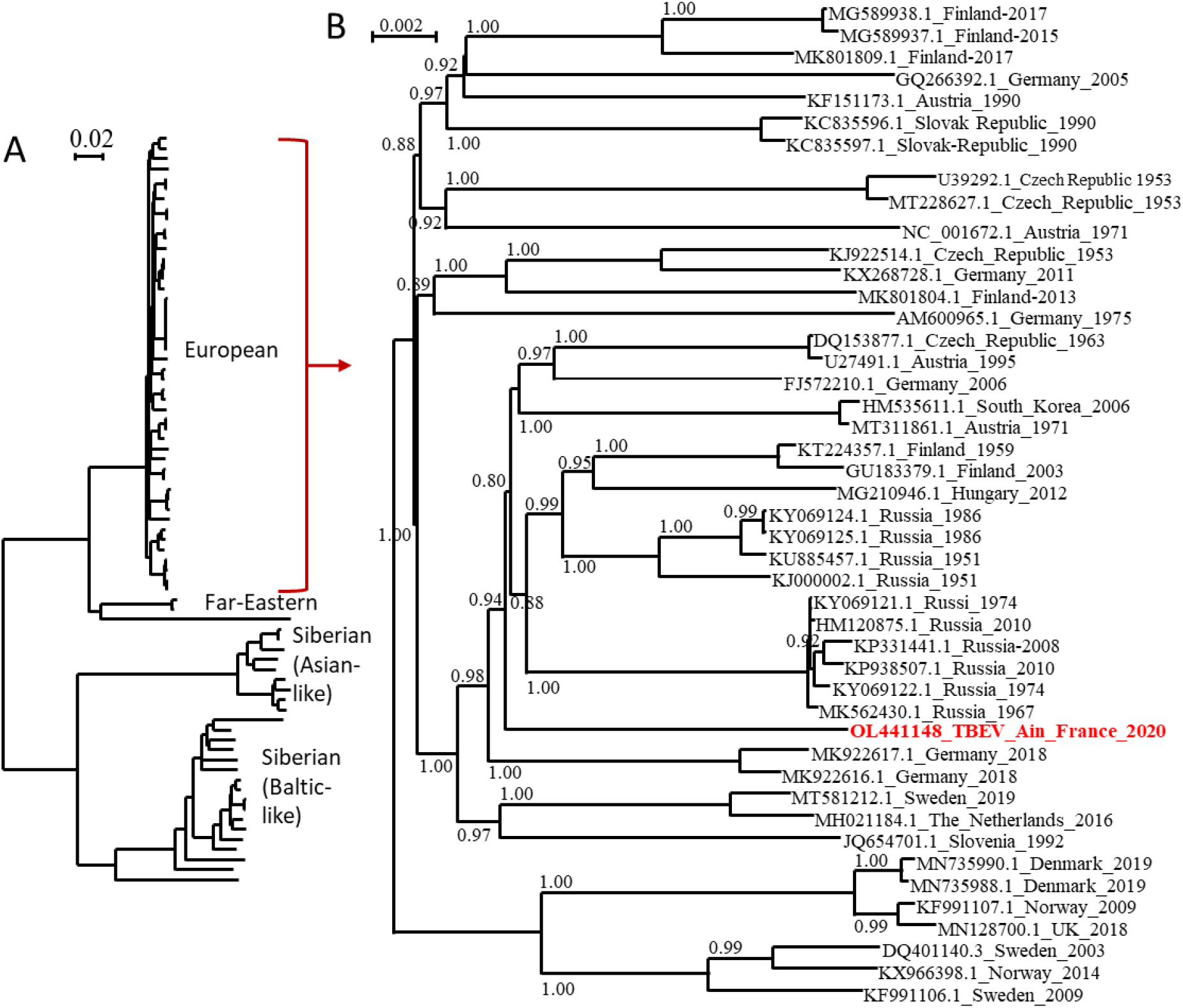
Phylogenetic tree of TBEV complete open reading frames available in GenBank and the TBEV-Ain-France-2020 obtained in the present study (marked red) using all sequences (A) or those corresponding to the European subtype only (B). The two trees were inferred in PhyML using the LG substitution model. Branch points indicate that results of Shimodaira-Hasgawa branch test > 0.8. Scale bar shows the number of nucleotide changes. Virus isolate names are given as follows: accession number from NCBI_ country name_year.

## Discussion

Food-borne TBE outbreaks had been reported in several central endemic European countries at a regular frequency, in particular in countries where the consumption of traditional raw milk products is popular (Kerlik, Avdičová et al. 2018). France is on the border of TBEV geographic range and cases resulting from tick bites are reported very occasionally. The first occurrence of TBE-food borne outbreak in France in 2020 has led to an unexpected high number of cases: 43 patients have developed clinical symptoms compatible with TBE, and all but one had consumed raw goat milk and/or cheese products originating from a single producer, while 41 cases could be confirmed (REFERENCE AUTRE PAPIER HUMAIN). A health alert was triggered within a month after the first patient showed symptoms and an in depth investigation of TBEV presence was conducted in the farm suspected to be the source of contamination and in animals in surrounding farms. We applied a “one health” approach, combining environmental, veterinary, virology and food microbiology approaches to identify the origin of infection and provide a range of epidemiological information of significance to public health decision makers. We characterized the TBEV strain circulating in Ain, detected viral genome presence in a batch of cheese, goat milk and in questing ticks, demonstrated recent infection of goats, assessed the enzootic hazard for TBEV exposure and we identified a wooded TBEV-focus area. This integrative investigation provides information that would allow the farmer and authorities to assess the risk of infection and develop control measures.

The initial investigation of seroprevalence (23%) in goat flock at the suspected farm reveals that the animals had been moderately exposed to the virus these past few years. In neighboring areas where TBEV is endemic such as Switzerland and Germany, seroprevalence varies from 1 to 80% with a median range values of 10-20% (calculated from (Klaus, Beer et al. 2012, Klaus, Ziegler et al. 2014, Rieille, Klaus et al. 2017, Casati Pagani, Frigerio Malossa et al. 2019)). Seroconversion results, presence of IgM antibodies and detection of the virus genome in milk indicate that infection was very recent for half of the animals that had been exposed to the virus (6 animals over 13). IgM were detected in one individual. Three individuals seroconverted after entering the stable, indicating that TBEV infection occurred a few days before they entered the stable. Indeed, seroconversion in goat and sheep occurs in 6 to 10 days (Van Tongeren 1955, Klaus, Ziegler et al. 2014, Paulsen, Stuen et al. 2019). One of these individuals, and two others excreted viruses in the milk 14 days after entering the stable. These results also support a recent infection given that virus excretion in goat and sheep milk is detected from 2 to 23 days after infection (Gresikova 1958, Gresíková, Sekeyová et al. 1975, Balogh, Ferenczi et al. 2010, Balogh, Egyed et al. 2012). So far, information is relatively scarce on the infection dynamics in ruminants, especially in naturally infected animals. Our investigation confirmed long-lasting excretion of the virus in milk in naturally infected goats since three individuals excreted the virus into milk for over 14 days post-infection. The date of the seroconversion of the additional seven TBEV-seropositive goats and two cows from the same farm is unclear. Previous studies suggest that TBEV-antibodies may persist up to 6 years, but the level of IgG produced decreases overtime (Klaus, Ziegler et al. 2014, Klaus, Ziegler et al. 2019). In agreement, even though the level of neutralizing antibodies remain high a year later, we observed a decrease in the level of IgG detected by ELISA. Thus, the high level of IgG as measured by ELISA competition indicates that the remaining animals have been likely infected or re-infected by the virus in 2020. Since no virus excretion have been observed in five experimentally pre-immunized goats (Balogh, Egyed et al. 2012) only naïve individuals would have excreted TBEV in milk. The high recent infection rate among naïve individuals of the flock would explain the high virus load detected in tank milk in June. Milk and cheese viral load was determined using a genome quantification assay, whereby milk sample concentration was estimated based on Ct threshold values derived from measurements of serial dilutions of infectious TBE viral particles with known concentrations (from 10^8^ to 10^3^ TCID50/mL). We evaluated a minimal detection of infectious viral particle at 10^4^ TCID50/mL of milk and 10^5^ TCID50/ mL after dissolution of 2.5g of cheese. In ticks, the mean viral copy number lies between 2 × 10 ^2^ to 4.8 x10^3^ RNA copies per sample (Liebig, Boelke et al. 2020, Liebig, Boelke et al. 2021). Our data confirmed the high virus load detected in tank milk. Moreover, the virus was detected in seven cheese (over 91 tested). Among them, one contaminated cheese was produced on April 28. This is outstanding especially when we consider that cheese were kept during several days at 1-5°C by consumers or producers before being analysed, which may have reduced persistence of TBEV load and hence its detection.

The high recent infection rate of the goat flock suggests regular contact between TBE-infected ticks and goats in spring 2020, when ticks started to be active. We found infected questing nymphs and adults in the wooded edge of the pasture (in the meadow between the pasture wood and the forest) and in the forest nearby. With a MIR of 0.22% (IC_95%_ : 0.03 – 0.80%) in nymphs and an infection rate of 1.8% (IC_95%_ : 0.1 – 9,9%) in adults, it appears that TBEV is well established in the forest near the pasture. Indeed, while the densities and prevalence of infected questing ticks may not be deemed exceptionally high compared to what is seen in endemic countries (Süss 2011), it is similar to the highest values observed in an endemic foci in Alsace region, East France (Bournez, Umhang et al. 2020). Since we captured few adult ticks (probably because the dragging method is not well suited to capture this stage), the density of infected adults was estimated with a low precision and might be higher than those observed in Alsace. We did not detect TBEV-RNA in small mammals trapped in the pasture, but we probably missed infected individuals since we only trapped few rodents. Indeed, the year 2020 was probably a low-phase of usual yearly-fluctuating rodent population. Overall, the results suggest that the presence of a large wooded area within the pasture frequently used by goats as a shelter as well as of a long pasture-forest edge where tick densities are relatively high have likely favoured the contacts between goats and TBEV-infected ticks. The forest may also represent a source of frequent introduction of infected ticks into the pasture through the movements of tick-feeding hosts.

This cluster surprised by its location: it occurred in the Ain department in France where TBEV human cases had never been reported. Our investigation in surrounding farms also revealed that TBEV was also present in various pastures surrounding the same single forest where we found infected ticks over at least two kilometers. In three of these farms, seroprevalence varied between 5.5% and 25%. With the exception of one individual (for which it was not possible to track back the pasture in which it had grazed), all seropositive animals had likely grazed within pasture adjacent to the same forest. These results indicate that the virus is well established in the area and probably in a larger extent in Ain department. Because of the absence of an active TBEV surveillance system assessing its distribution in France and potential misleading of TBE diagnosis, it is impossible to conclude whether the presence of TBEV in the department is ancient or whether it was recently introduced. In addition, diagnosis of human TBE cases was initially delayed as TBE serology was not systematically researched among patients presenting with meningitis or meningoencephalitis. Similarly, the presence of TBEV in centre France, in Loire and Haute-Loire departments, detected for the first time in 2017 and 2018 (Herpe, Schuffenecker et al. 2007, Velay, Solis et al. 2018) is impossible to date. Given the life-cycle of ticks with generally one meal per stage per year, we can only presume that TBEV-infected ticks have been present in the environment for at least one or two years before the occurrence of human cases. Evidence suggest that the geographic range of TBEV is currently expanding in Europe, especially in Western and Northern Europe (Csángó, Blakstad et al. 2004, Donoso Mantke, Escadafal et al. 2011, Rieille, Klaus et al. 2017, Andersen, Larsen et al. 2019, Casati Pagani, Frigerio Malossa et al. 2019, Smura, Tonteri et al. 2019, Alfano, Tagliapietra et al. 2020, Ullrich, Schranz et al. 2021) and this might have occurred in France as well. For instance, a recent expansion of TBEV in higher altitudes, other previously unaffected areas or ancient affected areas has been observed in several France’s neighbouring endemic countries, including Germany (Dobler, Erber et al. 2019, Hellenbrand, Kreusch et al. 2019), Switzerland (Rieille, Klaus et al. 2017, Casati Pagani, Frigerio Malossa et al. 2019, Desgrandchamps D. 2021) and Italy (Alfano, Tagliapietra et al. 2020). The virus was also detected for the first time in the UK in 2018 (Holding, Dowall et al. 2020) and in the Netherlands in 2015 (Jahfari, de Vries et al. 2017). TBEV expansion is believed to be driven by climate, landscape and anthropogenic changes (Randolph 2010, Jaenson, Hjertqvist et al. 2012, Holding, Dowall et al. 2020). Moreover, the first occurrence of TBEV in new areas in France since 2017 might be the results of exceptional high densities of infected ticks these past five years in endemic areas, which may have led to higher dispersal rates of infected ticks into new areas. Indeed, although TBE human incidence usually exhibit significant oscillations over time, in 2016-2020, it was persistently high in numerous endemic countries of Western and Central Europe (Germany, Switzerland, Czech Republic, Slovakia, Italy) including France (Dobler, Erber et al. 2019). This may partly be caused by high densities of infected ticks during those years. A high density of infected ticks during several years in endemic areas may increase the frequency of medium or long distance movements of infected ticks on their host (birds, cervids) and led to more successful introduction of the virus into new areas. Alternatively, it is also possible that TBEV has been circulating in Ain department, undetected, for decades and the 2020 outbreak could be the result of suitable conditions for its detection. Indeed, TBE cases are likely underdiagnosed in France given that human clinical cases are rare and local clinicians are unfamiliar with the disease (infection often asymptomatic, not yet notifiable disease in April 2020, low virus circulation, few TBEV foci, few human visits in infected areas) (Dollat, Bellanger et al. 2021). The year of 2020 was exceptional with a very high human incidence observed in numerous endemic European countries. In Switzerland and Germany, the notable increase of TBE cases observed in spring 2020 was partly associated to higher recreational activities in risky endemic areas around people home during the Covid-19 containment measures (Steffen, Lautenschlager et al. 2020, Ullrich, Schranz et al. 2021). In France, the Covid-19 containment measures may have also indirectly increase the consumption of local products and hence the emergence of a cluster of food-borne TBE cases in a restricted geographical area that was easier to detect. However, this was not the solely factor and an upsurge of infected ticks densities in 2020 likely have contributed to human incidence’s rise. Indeed, models forecasting the human TBE incidence based on demographic parameters, large-scale atmospheric circulation patterns, the fructification of the European beech (*Fagus sylvatica*) two years prior, and the national TBE vaccination coverage, already predicted an increase in TBE incidence in 2020 in Germany, Switzerland and Austria (Rubel and Brugger 2020, Rubel and Brugger 2021). In Germany, an unusually high density of infected adult ticks has also been observed in 2020 in some places (PROMED 2020).

We provide the first TBEV isolate responsible for as a source of dietary contamination in France, which will facilitate experimental studies. The genome sequences obtained were used to conduct phylogenetic analyses using the complete open reading frame. We revealed that the TBEV_Ain_France_2020 belonged to the European TBEV subtype but are distantly related to the strain circulating in the Alsace region and to other isolates from Europe, the closest relatives being isolates from Germany, Austria, Czech Republic, The Netherlands and Russia. The distinct genome of the French isolate would tend to indicate that this isolate has not recently spread to France, but has remained undetected and evolved locally for some time. Further studies in the department and neighbouring region are needed and are being set up to further characterize the focus-specific genetic diversity of TBEV and to confirm or disprove the ancient origin of this isolate, as well as the geographical extension.

In conclusion, the cluster of TBE alimentary cases in Ain was unexpected and surprised clinicians and local health authorities given the number of infected patients. The high recent infection rate of the goat flock probably led to a high virus load in tank milk in April and May, and facilitated infection by consumption of raw cheese. In addition, the Covid-19 containment measures in spring 2020 may have also indirectly increase the consumption of local products and hence the risk of food-borne TBE cases. The Ain outbreak also highlighted the need for improving surveillance, detection and prevention of TBE in France. Clearly, a better understanding of TBEV distribution in France, of local virus dynamic, and of food-borne contamination with a better identification of raw milk products being at risk is needed to better adapt surveillance and prevention measures. This is all more important that France produces each year 40,000 tonnes of raw milk-products and cheese and the local production and consumption of traditional delicacies is increasing. TBEV is now a notifiable disease in France, which will facilitate the systematic report of human cases and improve sensitizing medical communities for diagnostic in humans showing encephalitis in France. In endemic regions and those surrounding affected areas, patients with meningitis and encephalitis in the absence of other etiological diagnosis should to be more systemically screened for TBEV infection (Herpe, Schuffenecker et al. 2007, Velay, Solis et al. 2018, Botelho-Nevers, Gagneux-Brunon et al. 2019, Blanchon, Boulanger et al. 2020). Surveillance could also include serological testing for anti-TBEV antibodies in blood of animals or in milk tank to allow early detection of virus foci (Imhoff, Hagedorn et al. 2015, Rieille, Klaus et al. 2017, Wallenhammar, Lindqvist et al. 2020, Bauer, Könenkamp et al. 2021). Finally, in focus areas, TBEV virus vaccination in humans could be used to prevent tick bite or alimentary contamination (Chernokhaeva, Rogova et al. 2018, Kubinski, Beicht et al. 2020, Chitimia-Dobler, Lindau et al. 2021).

## Data Availability

All data produced in the present study are available upon reasonable request to the authors

## Acknowledgments

The authors would like to thank the clinicians of Bourg-en-Bresse and Oyonnax hospitals who were involved in patients care and initiated the health alert; in particular Dr Beaufils, Dr Canu, Dr Decouchon, and Dr Decroix.

