## Supplemental table for "A one health approach to investigating an outbreak of alimentary tick-borne encephalitis in a non-endemic area in France (Ain, Eastern France): a longitudinal serological study in livestock, detection in ticks, and the first TBE virus isolation and molecular characterization"

Table 1: Primers used for full-length genome sequencing of TBEV from milk and tick samples

| Name | Pool | Sequence | Size (nt) | %GC | Tm |
|---|---|---|---|---|---|
| scheme_1_LEFT | 1 | TGTTTCTACCAGTCGTGAACGT | 22 | 45.45 | 60.08 |
| scheme_1_RIGHT | 1 | CCCCAACAGAGTAATGACCAGC | 22 | 54.55 | 61.12 |
| scheme_2_LEFT | 2 | ATCAAAAGGACAGTGAGTGCCC | 22 | 50.00 | 61.00 |
| scheme_2_RIGHT | 2 | ATGGGATGGGATCAGCACTGAG | 22 | 54.55 | 62.07 |
| scheme_3_LEFT | 1 | ACGTGGATTGTTTTTGCCGGAA | 22 | 45.45 | 62.03 |
| scheme_3_RIGHT | 1 | GTAACACATCCACCCAGTTCCA | 22 | 50.00 | 60.41 |
| scheme_4_LEFT | 2 | CGTTGCACACACTTGGAAAACA | 22 | 45.45 | 60.85 |
| scheme_4_RIGHT | 2 | TTTGTTGGCGTCGTACACATGT | 22 | 45.45 | 61.49 |
| scheme_5_LEFT | 1 | TTGGAAAGGGTAGCATTGTGGC | 22 | 50.00 | 61.59 |
| scheme_5_RIGHT | 1 | CTGCGTTATTCCAGTTTTGCGC | 22 | 50.00 | 61.71 |
| scheme_6_LEFT | 2 | AACACCTTCCAACGGCTTGG | 20 | 55.00 | 61.12 |
| scheme_6_RIGHT | 2 | GGGATCCTACAGGGCTTTGTTC | 22 | 54.55 | 60.87 |
| scheme_7_LEFT | 1 | CACATGGAAGAGAGCTCCAACA | 22 | 50.00 | 60.47 |
| scheme_7_RIGHT | 1 | TGCTGTTGAAAGCACCACCAAG | 22 | 50.00 | 62.28 |
| scheme_8_LEFT | 2 | GACTGACAGTGATAGGAGAGCAC | 23 | 52.17 | 60.43 |
| scheme_8_RIGHT | 2 | GCTTCCCTCCTCAAATGTCTCC | 22 | 54.55 | 60.86 |
| scheme_9_LEFT | 1 | AGAGAGGTCTCAGAATGGTATGACA | 25 | 44.00 | 60.97 |
| scheme_9_RIGHT | 1 | CGAATTCTGCCACCGTGAAAAC | 22 | 50.00 | 61.09 |
| scheme_10_LEFT | 2 | TTTCATGGTGGGCACGGAAG | 20 | 55.00 | 60.91 |
| scheme_10_RIGHT | 2 | CACCTGTTCTGAATAACCGGGT | 22 | 50.00 | 60.47 |
| scheme_11_LEFT | 1 | CGTGGTGGACTCGGAGTTATTC | 22 | 54.55 | 60.91 |
| scheme_11_RIGHT | 1 | AAACAATGCCACTATTCCGGGG | 22 | 50.00 | 61.39 |
| scheme_12_LEFT | 2 | GAAATACGGCCAGTCCATGACC | 22 | 54.55 | 61.50 |
| scheme_12_RIGHT | 2 | CCCAATTCCAGGACCAGCAAAA | 22 | 50.00 | 61.53 |
| scheme_13_LEFT | 1 | TGCTCAGCGCATTTGCACT | 19 | 52.63 | 61.19 |
| scheme_13_RIGHT | 1 | CAAAACGCCAGCAGTCTGATTC | 22 | 50.00 | 60.84 |
| scheme_14_LEFT | 2 | AGAGAGCAGAAGGGATTGACCT | 22 | 50.00 | 60.75 |
| scheme_14_RIGHT | 2 | GCCAAAAAGCCATCATTCTCTCT | 23 | 43.48 | 59.81 |
| scheme_15_LEFT | 1 | GTGGAATGGCATCCGGAACTA | 21 | 52.38 | 60.24 |
| scheme_15_RIGHT | 1 | TGCCACATCGTGTGCAAGAC | 20 | 55.00 | 61.52 |
| scheme_16_LEFT | 2 | GTCTACAGGATTTTCAGCCCCG | 22 | 54.55 | 61.18 |
| scheme_16_RIGHT | 2 | TAGGCCATTTCCGTATAGCCCC | 22 | 54.55 | 61.87 |
| scheme_17_LEFT | 1 | GCTTGGGGCAATACCAATTGAT | 22 | 45.45 | 59.75 |
| scheme_17_RIGHT | 1 | TCCAGCCTGTTGGTCACTGA | 20 | 55.00 | 61.07 |
| scheme_18_LEFT | 2 | CTCCAACTCGTGTGGTACTCAA | 22 | 50.00 | 60.14 |
| scheme_18_RIGHT | 2 | CGCCCTTCATACTCAGTGATCC | 22 | 54.55 | 60.73 |
| scheme_19_LEFT | 1 | CTGGTCTTGATGACAGCGACAC | 22 | 54.55 | 61.67 |
| scheme_19_RIGHT | 1 | TGAGCTCGACCTTCCCATCAA | 21 | 52.38 | 61.28 |
| scheme_20_LEFT | 2 | ATATCTCGGAGATGGGAGCCAA | 22 | 50.00 | 60.68 |
| scheme_20_RIGHT | 2 | CCAGCCACGGTGTAAAGTCA | 20 | 55.00 | 60.27 |
| scheme_21_LEFT | 1 | CGGTCACTTTCGACTCACTGAA | 22 | 50.00 | 60.72 |
| scheme_21_RIGHT | 1 | TGCATGAGTGTGTAGAAGACATCC | 24 | 45.83 | 60.94 |
| scheme_22_LEFT | 2 | GTTCTGACAGGAATGTCGGGAG | 22 | 54.55 | 60.85 |
| scheme_22_RIGHT | 2 | ATGCCAGTTTGTTGTCGTCACT | 22 | 45.45 | 61.19 |

| | | | | | |
|---|---|---|---|---|---|
| scheme_23_LEFT | 1 | CCGGAGTGGCTCTCATCTTCTA | 22 | 54.55 | 61.19 |
| scheme_23_RIGHT | 1 | ATGTCCCGCCACACCAAAGA | 20 | 55.00 | 62.08 |
| scheme_24_LEFT | 2 | ATCCAACAACTTGTCAACAGCG | 22 | 45.45 | 60.14 |
| scheme_24_RIGHT | 2 | TCTGTTATGGAGGCCACCGTTC | 22 | 54.55 | 62.50 |
| scheme_25_LEFT | 1 | AAAGGAAAATGAGTCTGGTGTTGG | 24 | 41.67 | 59.98 |
| scheme_25_RIGHT | 1 | TGGTCTCTCCTCTTCTGAGCAA | 22 | 50.00 | 60.68 |
| scheme_26_LEFT | 2 | ACTGCACCAGGGAGGAATTCTT | 22 | 50.00 | 61.82 |
| scheme_26_RIGHT | 2 | ATCTGGGCTGCTCTCTCCAAT | 21 | 52.38 | 61.08 |
| scheme_27_LEFT | 1 | AGATCAGGAATGGACGTGTTCAG | 23 | 47.83 | 60.62 |
| scheme_27_RIGHT | 1 | TCCCAGGTCAAGTTCAGGCA | 20 | 55.00 | 61.08 |
| scheme_28_LEFT | 2 | GTCAACGTACAGTCGAGGAAACT | 23 | 47.83 | 60.80 |
| scheme_28_RIGHT | 2 | TCCTGTGCCTTTGTGTCAACTT | 22 | 45.45 | 60.80 |
| scheme_29_LEFT | 1 | GTGCGCATGGCTATGACTGA | 20 | 55.00 | 60.83 |
| scheme_29_RIGHT | 1 | ACTCCGAACTCTCCCAGTTTCT | 22 | 50.00 | 60.94 |
| scheme_30_LEFT | 2 | GAGAGAGAAAGGCACCTCATGG | 22 | 54.55 | 60.60 |
| scheme_30_RIGHT | 2 | GCTTTTTGCATTATTGTGGTTGCC | 24 | 41.67 | 60.93 |
| scheme_31_LEFT | 1 | ACCAATGCAGACTTAGAGGATGAA | 24 | 41.67 | 60.10 |
| scheme_31_RIGHT | 1 | CAAATCTGTCATCCAAGGGCCT | 22 | 50.00 | 60.81 |
| scheme_32_LEFT | 2 | GCTGGCTGAAAGAACATGGAGA | 22 | 50.00 | 61.06 |
| scheme_32_RIGHT | 2 | CTGAGTTAATGGCAAGCCCGAG | 22 | 54.55 | 61.75 |
| scheme_33_LEFT | 1 | GAGACGGCCTGCCTTTCAAA | 20 | 55.00 | 60.91 |
| scheme_33_RIGHT | 1 | TCCATACAGGAGAGATAGTCCTTGA | 25 | 44.00 | 60.26 |
| scheme_34_LEFT | 2 | AGAATGGGCCAAGAACATCTGG | 22 | 50.00 | 60.81 |
| scheme_34_RIGHT | 2 | CCATGATCTGTGGCTTCGCTTC | 22 | 54.55 | 62.06 |
| scheme_35_LEFT | 1 | CACTACGGGACTGCTTCATAGC | 22 | 54.55 | 60.98 |
| scheme_35_RIGHT | 1 | AGGAGGAAAAATCCTGAAGAGAGC | 24 | 45.83 | 60.65 |

Table 2: Sample barcoding performed for full-length virus genome sequencing

| Sample | Barcode Pool 1 | Barcode Pool 2 |
|---|---|---|
| Infected Ticks pool 1 | NB01 | NB08 |
| Infected Ticks pool 2 | NB02 | NB09 |
| Infected Ticks pool 3 | NB03 | NB10 |
| Contaminated milk 1 | NB04 | NB11 |
| Contaminated milk 2 | NB05 | NB12 |
| Contaminated milk 3 | NB06 | NB13 |
| Contaminated milk 4 | NB07 | NB14 |

Table 3 :

| Primer name | Sequence | |
|---|---|---|
| | Forward | Reverse |
| **TBEV_1** | AGACAGCTTAGGAGAACAAGAGCT | GCAATCGTCATCCCCAACAGAG |
| **TBEV_2** | AGGACAGTGAGTGCCCTAATGG | ATGGGATGGGATCAGCACTGAG |
| **TBEV_3** | ACGTGGATTGTTTTTGCCGGAA | AAGGCTTCCCCTCAGCTGTTAT |
| **TBEV_4** | TGCACACATTTGGAAAACAGGGA | TTTTTGCCTCACAAGCCACCTT |
| **TBEV_5** | GGTGGTACAGTGTGCAAGAGAG | TCCCTCATGTTTCCATGGCAGA |
| **TBEV_6** | AGACCGTCATCCTTGAGCTTGA | TGACCACTGTATCATGCCCACT |
| **TBEV_7** | CGAAGTGGGACTGGAAAAACTGA | GTGCTCCCCTATCACTGTCAGT |
| **TBEV_8** | GTTGGGGAACTGAGTTATCAATGGT | TTCTGAGACCTCTCTCCACACG |
| **TBEV_9** | GTTTTGGCCATGACCCTTGGAG | GCTCCAGATCATTGAATGGCCC |
| **TBEV_10** | GTGGACAAGTTTGACCCCACTG | GAGCAGTTCCTCAGGTCAGTGA |
| **TBEV_11** | GCATGGCAATCCACACAGATCA | GCAGCACCATTCTGGGATAACC |
| **TBEV_12** | ACGCCTATCCGAGTCATCAGAG | TGACAAGCAAAGCGAGAACGAC |
| **TBEV_13** | CCCCGGAATAGTGGCATTGTTT | CCGTGCAAGCCCTGAATATCAG |
| **TBEV_14** | TTTTGCTGGTCCTGGAATTGGG | AGCATGACCCCTACCACAGTTA |
| **TBEV_15** | AGAGGGCTCTGGAATCAGACTG | CACACCAAGAATGCCTGACCAA |
| **TBEV_16** | ACTTTCACTTGACTGAGCTCGAG | TTTTCCTCCAGACTCCAGGCTC |
| **TBEV_17** | GGCGCTGTCTATTGATGATGCT | GCATGTCCAGCACTGTGATCTG |
| **TBEV_18** | ATGAGACCTACGTCAGCAGCAT | TATGCTGTGAGGGTCAGTCCAG |
| **TBEV_19** | CGCAACCTATGTCAACAGACGG | CAGGCTTCTCATCCCTCACTCT |
| **TBEV_20** | TAGCTCGCACCTTGAGACAGAA | CTGGTCCATAAAAGGTGGCCAC |
| **TBEV_21** | ACAGTGTGATGATGATGACAGTGG | ACGCCACGAACTCTTTGATGTC |
| **TBEV_22** | GATCGAAGCTGGACATGGGAAG | ATACGGCTGATTGAAGTCCGGA |
| **TBEV_23** | GAGGCCTTTCTGACCATGGTTG | CATTCGTCCATTCACTCCACGG |
| **TBEV_24** | ACTGGTTGCAGCCAATGAGATG | TGTCAATTCAGCCTCCAGACCA |
| **TBEV_25** | GGTGGTGTCACTGATTGGAGCT | TACCCCTGACCACACCACTCAT |
| **TBEV_26** | GAACGGTGGCCTCCATAACAGA | CCGAGAGACAGCCAATCCCATA |
| **TBEV_27** | ATTCTTTGTGTACAGGCGCACT | TGCTCTCTCCGATGTCACACAT |
| **TBEV_28** | AACAAGCCTGGGTTGGAACTTG | AACCGAGCCAAAAGTTTCCTCG |
| **TBEV_29** | AGGAACTCCACCCATGAGATGT | AAAGCCGTTGTGTCAGTCATGG |
| **TBEV_30** | GCGTCACTGATCAATGGGGTTG | CCCATGAGGTGCCTTTCTCTCT |
| **TBEV_31** | GATGAGCAAAACAGATGGGCGA | GTAACTTTCGTGTCCCAGCCAG |
| **TBEV_32** | AAGCTTGAACTACCTGGGCTGG | TCTATGACCCCTTCCCCTTCCA |
| **TBEV_33** | AGAGATCAAAGAGGTTCGGGCC | GTTCATCTTGGTCTCTGCACGG |
| **TBEV_34** | CTTATCAAGCTGGGAGGAAGTCC | CATCCCTCCACTCCATGACCTT |
| **TBEV_35** | TGACCACAGAAGACATGCTGGA | ACACTCTGTGAGTTGCTTGCTT |
| **TBEV_36** | TGGAGAGCTCAATAATCTAAACCCAGA | GTGGCTCAGGGAGAACAAGAAC |
